# Immunogenicity of a Two Dose Regimen of Moderna mRNA Beta/Omicron BA.1 Bivalent Variant Vaccine Boost in a Randomized Clinical Trial

**DOI:** 10.1101/2023.06.06.23290973

**Authors:** Nadine G. Rouphael, Angela R. Branche, David J. Diemert, Ann R. Falsey, Cecilia Losada, Lindsey R. Baden, Sharon E. Frey, Jennifer A. Whitaker, Susan J. Little, Satoshi Kamidani, Emmanuel B. Walter, Richard M. Novak, Richard Rupp, Lisa A. Jackson, Tara M. Babu, Angelica C. Kottkamp, Anne F. Luetkemeyer, Lilly C. Immergluck, Rachel M. Presti, Martín Bäcker, Patricia L. Winokur, Siham M. Mahgoub, Paul A. Goepfert, Dahlene N. Fusco, Robert L. Atmar, Christine M. Posavad, Antonia Netzl, Derek J. Smith, Kalyani Telu, Jinjian Mu, Mat Makowski, Mamodikoe K. Makhene, Sonja Crandon, David C. Montefiori, Paul C. Roberts, John H. Beigel, the COVAIL Study Team

## Abstract

In this brief report, we compare the magnitude and durability of the serologic response of one versus two doses (separated by 56 days) of a variant vaccine (Moderna mRNA-1273 Beta/Omicron BA.1 bivalent vaccine) in adults.

## Introduction

A single boost with currently authorized bivalent prototype/Omicron BA.4/5 vaccines leads to a serological advantage compared to ancestral vaccine boosting against Omicron subvariants.^1^ Although additional doses of variant boosters are recommended for individuals at risk for severe disease^2^, it is unclear whether two doses of variant vaccines further boosts humoral immunity to SARS-CoV-2 variants.

## Methods

Adult participants were randomized 1:1 to receive one or two doses (separated by 56 days) of Moderna mRNA-1273 Beta/Omicron BA.1 bivalent vaccine (50 mcg) ≥16 weeks from last monovalent prototype vaccination. Beta/Omicron BA.1 was chosen to cover an antigenic space distant to D614G from which new variants may emerge. We assessed pseudovirus neutralization titers^3^ at 14, 28, 90 and 180 days after the last vaccination against D614G, Omicron BA.1, Omicron BA.4/5, B.1.351 and B.1.617.2. Participants were removed from analysis following self-reported infection, N-antibody positivity or receipt of an out-of-study booster.

## Results

One hundred participants were enrolled in the one-dose and 102 in the two-dose arms. In previously uninfected participants, Day 29 ID_50_ GMTs in the one-dose arm against D614G, BA.1, BA.4/5, B.1.351 and B.1.617.2 were 12,609, 2,824, 1,219, 7,075 and 7,024, respectively (Figure 1, Tables S1/S2). Corresponding ID_50_ GMTs in the two-dose arm, 28 days after last dose, were 12,464, 2,891, 1,391, 7,507 and 7,708, respectively. To compare the degree of antibody waning, the Geometric Mean Fold Drop (GMFD) from Day 29 to 180 days after final boosting were calculated; for the one-dose/two-dose arms GFMD were 4.3/3.0 for D614G, 4.8/4.3 for BA.1, 4.6/3.3 for BA.4/5, 5.5/4.0 for B.1.351 and 4.5/3.5 for B.1.617. At enrollment, 18% and 21% of the one and two-dose arms, respectively, had a prior SARS-CoV-2 infection by self-report or N-antibody testing (Table S7). Titers were higher in these previously infected participants at all timepoints with similar findings following one or two doses (Tables S3 and S4).

**FIGURE 1.**
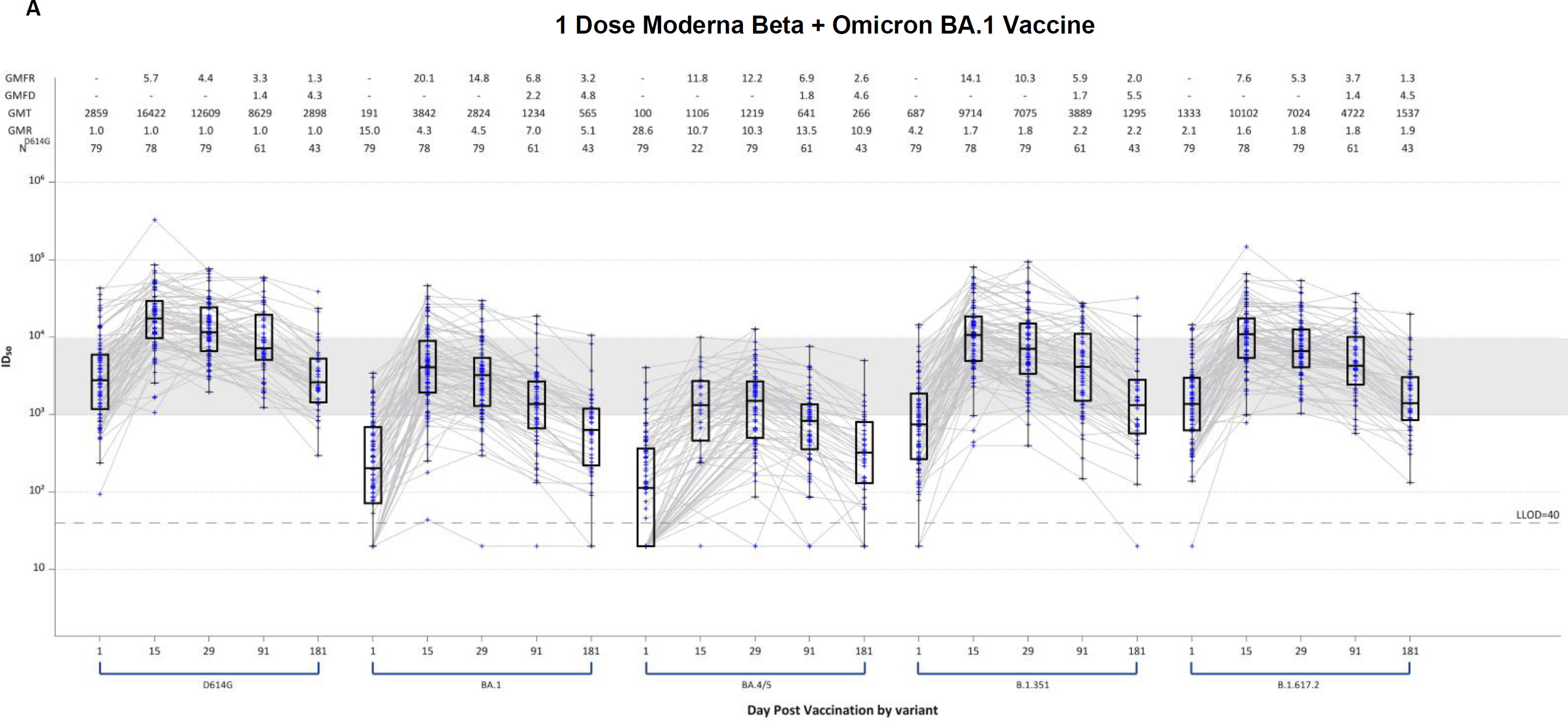
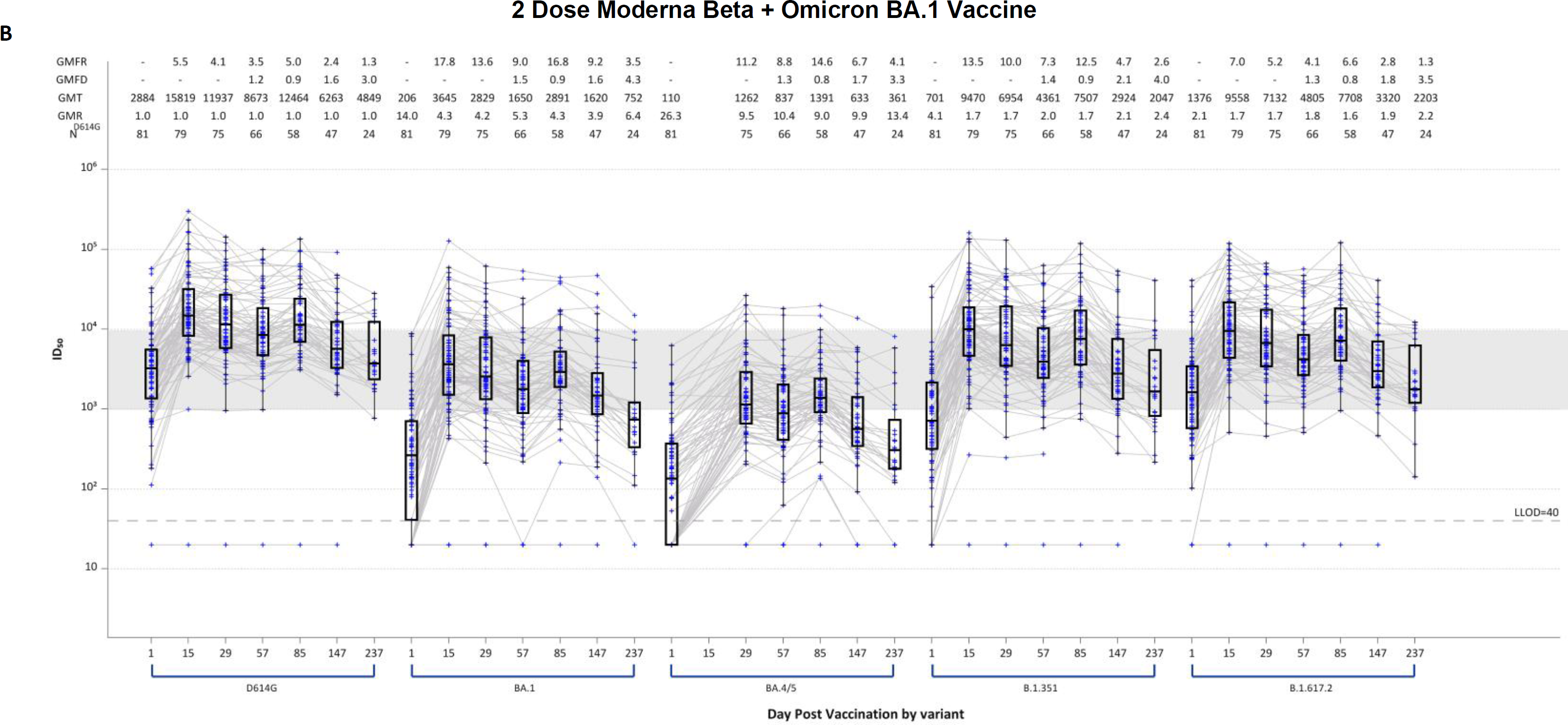
Pseudovirus Neutralization ID_50_ Titers by Timepoint (baseline, Day 15 and 29 for both arms and 90 and 180 days after Vaccination for each arm, respectively and Day 85 for the 2 dose sample prior to receipt of second dose) and Variants, (D614G, Omicron BA.1 [B.1.1.529], BA.4/BA.5, Beta [B.1.351] and Delta [B.1.617.2]) before and after vaccination with 1 dose of 50 mcg of Moderna mRNA-1273 Beta/Omicron BA.1 (A) or 2 doses (given 56 days apart) of 50 mcg Moderna mRNA-1273 Beta/Omicron BA.4/5 (B) in participants previously uninfected at baseline. Boxes with horizontal bars denote interquartile range (IQR) and median ID_50_, respectively. Whisker denotes 95% confidence interval. GMFR, geometric mean fold rise from baseline. GMT, geometric mean titer. LLOD, lower limit of detection of the assay.

## Discussion

A two-dose boosting regimen with a variant vaccine, albeit given 2 months apart, did not increase serological responses compared to a single variant vaccine boost, contrary to what has been observed after AS03-adjuvanted H5N1 influenza virus immunization.^4^ However, durability of the responses appears to be slightly improved with two doses. It is possible that a boost with more contemporary variant vaccines, or two doses given with a longer interval, could lead to different results. This study is the first in humans to show serological responses after a two-dose boost with a SARS-CoV-2 variant vaccine.

## Data Availability

All data produced in the present study are available upon reasonable request to the authors

## Acknowledgments

We would like to thank all participants who contributed to the study and the NIAID SARS-CoV-2 Assessment of Viral Evolution (SAVE) program team for their consultation regarding the study arm design and variant vaccine selection.

## Data availability

All data produced in the present study are available upon reasonable request to the authors.

## RLA, TMB, SMM, MB, MKM, JHB, SC, ACK, CL, PCR, RR, JAW

## SUPPLEMENTAL MATERIALS

### Study Design and Eligibility Criteria

This phase 2 open-label, randomized, clinical trial was performed at 22 sites in the US (Table S5) enrolling all participants from March to May 2022. Eligible participants were healthy adults (with or without a history of prior SARS-CoV-2 infection) who had received a primary series and a single homologous or heterologous boost with an approved or emergency use authorized prototype COVID-19 vaccine (Table S6). The most recent vaccine dose, and/or prior infection must have occurred at least 16 weeks prior to randomization. Full eligibility criteria are described at clinicaltrials.gov (NCT 05289037).

Two hundred and two eligible participants were stratified by history of confirmed SARS-CoV-2 infection, and randomly assigned for each arm in an equal ratio.

After informed consent, participants underwent screening, including confirmation of COVID-19 vaccination, medical history, a targeted physical examination, and a urine pregnancy test (if indicated). Samples were collected at Day 1, Days 15, 29, 57 (2-dose group only), and 85 (2-dose group only) and 3, 6, 9 and 12 months after last dose of vaccine. Immunologic data are currently available up to Day 181 for the 1-dose arm and through Day 237 (180 dose after second dose) for the 2-dose arm.

The trial was reviewed and approved by a central institutional review board and overseen by an independent Data and Safety Monitoring Board. Participants provided written informed consent before undergoing trial-related activities. The trial was sponsored and funded by the National Institutes of Health (NIH). The National Institute of Allergy and Infectious Diseases NIAID SARS-CoV-2 Assessment of Viral Evolution (SAVE) program team was consulted to inform study arm design and variant vaccine selection.

### Trial vaccine

The Beta/Omicron BA.1 bivalent trial vaccine was provided by Moderna (total amount of 50 mcg per vaccine; 25 mcg of each component). The vaccine candidates are manufactured similarly to their corresponding authorized or approved vaccines in the US or Europe.

### Study outcomes

The primary objective was to evaluate humoral immune responses of candidate SARS-CoV-2 variant vaccines.

### Immunogenicity assays

SARS-CoV-2 neutralization titers, expressed as the serum inhibitory dilution required for 50% neutralization (ID_50_), were assessed using pseudotyped lentiviruses^1^ presenting SARS-CoV-2 spike mutations tested in the Monogram laboratory (San Francisco, CA) lab for the following strains: D614G, Omicron BA.1, Omicron BA.4/5, B.1.351 and B.1.617.2.

### Pre-specified immunogenicity endpoints Statistical analysis

The primary objective of this study is to evaluate the magnitude and breadth of SARS-CoV-2 specific antibody titers in serum samples by estimating 95% confidence intervals (CI) for the geometric mean titer (GMT) at each timepoint when samples are collected. No formal hypothesis tests were planned.

The geometric mean fold rise (GMFR) is calculated as the geometric mean of titers at a timepoint divided by titers at Day 1. The geometric mean ratio to D614G (GMR_D614G_) is the geometric mean of the ratio of titers for a variant of concern to titers against D614G.

The GMFD was calculated by dividing the result at Day 29 by the result at each day after Day 29 and then calculating the geometric mean. Seropositive rate is calculated as the proportion of participants with titers above the lower limit of detection (LLOD). 95% CI for GMT, GMFR, GMFD and GMR_D614G_ are calculated using the Student’s t-distribution and 95% CI for seropositive rate is calculated using the Clopper-Pearson binomial method. For the purpose of analysis, participants were defined as previously infected by positive N antibody or self-report of a confirmed positive antigen or PCR testing. Participants were removed from analysis at timepoints following self-reported infection, N-antibody positivity or receipt of an out-of-study booster. Participants randomized to the 2-dose arm that did not receive the second dose were removed from the analysis after Day 29. This consisted of 11 participants who tested positive for COVID-19 prior to receiving the second dose, three participants who had adverse events and one participant who withdrew from the two-dose arm due to reactogenicity experienced after the first dose.

## COVAIL Study Team Authors

**George Washington University, Washington D.C**

David J. Diemert, MD; Elissa Malkin, DO; Jeffrey M. Bethony, PhD; Aimee Desrosiers, PA-C; Marc Siegel, MD

**University of Rochester VTEU, Rochester, NY**

Angela R. Branche, MD; Ann R. Falsey, MD; Edward Walsh, MD; Patrick Kingsley, BS; Michael Peasley, BS

**Emory University Hope Clinic, Decatur, GA**

Nadine G. Rouphael, MD; Cecilia Losada, MD; Daniel S. Graciaa, MD; Hady Samaha, MD; Cassie Grimsley Ackerley, MD; Kristen E. Unterberger, PA

**Brigham and Women’s Hospital, Harvard Medical School, Boston, MA**

Lindsey R. Baden, MD; Amy C. Sherman, MD; Stephen R. Walsh, MD; Alexandra Tong, BS; Rebecca Rooks, BS

**Saint Louis University, St. Louis, MO**

Sharon E. Frey, MD; Getahun Abate, MD, PhD; Zacharoula Oikonomopoulou, MD; Daniel F. Hoft, MD, PhD; Irene Graham, MD

**Departments of Molecular Virology and Microbiology and Medicine, Baylor College of Medicine, Houston, TX**

Jennifer A. Whitaker, MD; Hana M. El Sahly, MD; Wendy A. Keitel, MD; C. Mary Healy, MD

**Department of Medicine, Division of Infectious Diseases and Global Public Health, University of California San Diego, La Jolla, CA**

Susan J. Little, MD; Thomas C.S. Martin, MD; Nicole Carter, MPH; Steven Hendrickx, RN

**Center for Childhood Infections and Vaccines (CCIV) of Children’s Healthcare of Atlanta and Emory University Department of Pediatrics, Atlanta, GA**

Evan J. Anderson, MD; Christina A. Rostad, MD; Satoshi Kamidani, MD; Etza Peters, RN

**Duke Human Vaccine Institute, Duke University School of Medicine, Durham, NC**

Emmanuel B. Walter, MD, MPH; Michael J. Smith, MD, MSCE; M. Anthony Moody, MD; Kenneth E. Schmader, MD

**University of Illinois at Chicago-Project WISH, Chicago, IL**

Richard M. Novak, MD; Benjamin G. Ladner, MD; Andrea Wendrow, RPh; Jessica Herrick, MD

**University of Texas Medical Branch, League City, TX**

Richard Rupp, MD; Laura Porterfield, MD

**Kaiser Permanente Washington Health Research Institute, Seattle, WA**

Lisa A. Jackson, MD, MPH; Maya Dunstan, MS, RN; Rebecca Lau, PharmD; Barbara Carste, MPH

**Departments of Medicine, Epidemiology, and Laboratory Medicine & Pathology, University of Washington, Vaccines and Infectious Diseases Division, Fred Hutchinson Cancer Center, Seattle, WA**

Tara M. Babu, MD, MSCI; Anna Wald, MD, MPH; Taylor Krause, BA; Kirsten Hauge, MPH

**NYU VTEU Manhattan Research Clinic at NYU Grossman School of Medicine, New York, NY**

Angelica C. Kottkamp, MD; Mark J. Mulligan, MD; Tamia Davis, NP; Celia Engelson, NP; Vijaya Soma, MD

**Zuckerberg San Francisco General, University of California San Francisco, San Francisco, CA**

Anne F. Luetkemeyer, MD; Chloe Harris, BA; Azquena Munoz Lopez, BS

**Morehouse School of Medicine, Atlanta, GA**

Lilly C. Immergluck, MD; Erica Johnson, PhD; Austin Chan, MD

**Washington University School of Medicine, St. Louis, MO**

Rachel M. Presti, MD, PhD; Jane A. O’Halloran, MD, PhD; Ryley M. Thompson

**NYU VTEU Long Island Research Clinic at NYU Long Island School of Medicine, Mineola, NY**

Martín Bäcker, MD; Kimberly Byrnes, RN; Asif Noor, MD

**University of Iowa College of Medicine, Iowa City, IA**

Patricia L. Winokur, MD; Jeffery Meier, MD; Jack Stapleton, MD

**Howard University College of Medicine, Howard University Hospital, Washington D.C**

Siham M. Mahgoub, MD; Celia Maxwell, MD; Sarah Shami, PharmD

**University of Alabama at Birmingham, Birmingham, AL**

Paul A. Goepfert, MD

**Tulane University School of Medicine, New Orleans, LA**

Dahlene N. Fusco, MD; Arnaud C. Drouin, MD; Florice K. Numbi, MD

**University of Maryland**

Kirsten E. Lyke, MD

**IDCRC Principal Investigators**

David S. Stephens, MD; Kathleen M. Neuzil, MD

**IDCRC Leadership Operations Center**

Monica M. Farley, MD; Jeanne Marrazzo, MD; Sidnee Paschal Young

**IDCRC Clinical Operations Unit**

Jeffery Lennox, MD; Robert L. Atmar, MD; Linda McNeil FHI360

**IDCRC Statistical and Data Science Unit**

Elizabeth Brown, PhD

**IDCRC Laboratory Operations Unit – Fred Hutchinson Cancer Center, Seattle, WA**

Christine M. Posavad, PhD; Megan A. Meagher, BS; Julie McElrath, MD; Mike Gale, PhD

**FHI360, Durham, NC**

Kuleni Abebe, MSc

**The Emmes Company, LLC, Rockville, MD**

Mat Makowski, PhD; Heather Hill, MS; Jim Albert, MS; Holly Baughman; Lisa McQuarrie, MS; Kalyani Telu, MS; Jinjian Mu, PhD

**Clinical Monitoring Research Program Directorate, Frederick National Laboratory for Cancer Research, Frederick, MD**

Teri C. Lewis, BS; Lisa A. Giebeig, MS; Theresa M. Engel, MFS; Caleb J. Griffith, MPH; Wendi

L. McDonald, BSN; Alissa E. Burkey, MS; Lisa B. Hoopengardner, MS; Jessica E. Linton, MS; Nikki L. Gettinger, MPH

**Department of Surgery and Duke Human Vaccine Institute, Duke University School of Medicine, Durham, NC**

David C. Montefiori, PhD; Amanda Eaton, MBA

**Smith’s Laboratory, Cambridge, UK**

Derek J. Smith, PhD; Antonia Netzl; Samuel H. Wilks, PhD; Sina Türeli, PhD

**Division of Microbiology and Infectious Diseases, National Institute of Allergy and Infectious Diseases, National Institutes of Health, Bethesda, MD**

Mamodikoe Makhene, MD; Mohamed Elsafy, MD; Rhonda Pikaart-Tautges, BS; Janice Arega, MS; Binh Hoang, RPh; Dan Curtin; Hyung Koo, BSN; Elisa Sindall, BSN; Sonja Crandon, BSN; Marciela M. DeGrace, PhD; Diane J. Post, PhD; Paul C. Roberts, PhD; John H. Beigel, MD

## COVAIL Manuscript Study Members

**Emory University Hope Clinic, Decatur, GA**

Nadine G. Rouphael, MD; Cecilia Losada, MD; Daniel S. Graciaa, MD; Hady Samaha, MD; Cassie Grimsley Ackerley, MD; Kristen E. Unterberger, PA; Amy Anderson, BSN; Mary Atha, ACNP; Kareem Bechnak, BSN; Sarah Bechnak, BSN; Mary Bower, BSN; Laura Clegg, RN; Matthew Collins, MD, PhD; Francine Dyer, RN; Srilatha Edupuganti, MD; Rebecca Fineman, BS; Tigisty Girmay, MSN; Rebecca Gonzalez, PharmD; Natalie Gray, BS; Evan Gutter, MPH; Lisa Harewood; Chris Huerta, MSc; Brandi Johnson, BS; Lauren Johnson, MPH; Colleen Kelley, MD; Alexandra Koumanelis, BA; Deborah Laryea, BSN; Hollie Macenczak, BSN; Nour Makkaoui, MD; Michele McCullough, MPH; Tuong-Vy Ngo, PharmD; Eileen Osinski, BS; Julia Paine, BS; Bernadine Panganiban, BS; Rose Pope, RN; Paulina Rebolledo, MD; Susan Rogers, RPh; Erin Scherer, PhD; Veronica Smith, NP-C; Andre Stringer, BS; Jessica Traenkner, PA; Dongli Wang, BS; Alahna Watson, BA; Stacey Wheeler, RN; Jean Winter; Jianguo Xu, PhD

**Brigham and Women’s Hospital, Harvard Medical School, Boston, MA**

Lindsey R. Baden, MD; Amy C. Sherman, MD; Stephen R. Walsh, MD; Alexandra Tong, BS; Rebecca Rooks, BS; Jane A. Kleinjan, NP; Jon A. Gothing, NP; Andres A. Avila Paz, BA; Muneerah M. Aleissa, PharmD, MPH; Bethany Evans, BA; August Heithoff, BS; Natalie E. Izaguirre, MS; Hannah Jin, MPH; Urwah Kanwal, BS; Austin Kim, BS; Julia E. Klopfer, BS; Christina Montesano, BS; John Almeida, BA; Emily S. Koleske, BS; Hannah Levine, BS; Nicholas P. Morreale, BS; Omolola Ometoruwa, BS; Jun Bai Park Chang, BS; Anna F. Piermattei, BA; Djenane M. Pierre, BS; Megan Powell, BA; Kevin Zinchuk, PharmD; Stephanie Pickford, PharmD; Charles M. Kelly III, PharmD; Xiaofang Li, PhD; John Kupelian, BS; Kimberly Dufresne, BS; Xiaoguang Fan, MD, PhD; Xi Zhang, PhD; Esther Arbona-Haddad, MD; Jose Humberto Licona, MD

Evan J. Anderson, MD; Christina A. Rostad, MD; Satoshi Kamidani, MD; Etza Peters, RN; Larry Anderson, MD; Julia Bartol; Leisa Bower, RN; Natsuko Campbell, RN; Lisa Harewood; Hui-Mien Hsiao; Laila Hussaini, MPH; Inara Jooma; Gidget Kettle, RN; Marcia Lewis, RN; Wensheng Li; Cindy Lubbers, RN; Lisa Macoy, RN; Molly Morrison, Heather Nurse, RN; Anna Siaw-Anim; Kathleen Stephens, RN; Madeline Taylor; Ashley Tippett, MPH; Lauren Nolan, PA

**Zuckerberg San Francisco General, University of California San Francisco, San Francisco, CA**

Anne F. Luetkemeyer, MD; Chloe Harris, BA; Azquena Munoz Lopez, BS; Daniel Berrner; Dennis Dentoni-Lasofsky, MSN; John Dwyer, RN; Suzanne Hendler, BSN; Elvira Gomez, MPH; WeyLing Phuah, PharmD; Jaime Velasco, BA; Veronica Viar, MS

**George Washington University, Washington D.C**

David J. Diemert, MD; Elissa Malkin, DO; Jeffrey M. Bethony, PhD; Aimee Desrosiers, PA-C; Marc Siegel, MD; Nikita Schroll-McLaughlin, MS; Jonathan Manning, BA; Jane Ryu, MS; Hanna-Grace Rabanes, MPH; Khadija Khan, MPH; Laura Vasquez, MPH; Caroline Thoreson, PA-C; Larissa Scholte, PhD; Rafaela Thur, DVM; Peyton St. John, BS; Dorinne Mettle-Amuah, PharmD

**University of Iowa College of Medicine, Iowa City, IA**

Patricia L. Winokur, MD; Jeffery Meier, MD; Jack Stapleton, MD; Laura Stulken, PA; Theresa Hegmann, PA; Deb Pfab, RN; Elizabeth Morgan, RN; Susan Herman, RN; Angel Peguero, CMA; Michelle Rodenburg; Alfred J. Carr; Delilah Johnson

**Washington University School of Medicine, St. Louis, MO**

Rachel M. Presti, MD, PhD; Jane A. O’Halloran, MD, PhD; Michael Klebert, RN, PhD; Ryley M. Thompson; Alem Haile; Kim Gray, NP; Chapelle Ayres; Delaney Carani, RN; Michael Royal; John Tran; Laura Blair; Anita Afghanzada; Natalie Schodl

**NYU VTEU Manhattan Research Clinic at NYU Grossman School of Medicine, New York, NY**

Angelica C. Kottkamp, MD; Tamia Davis, NP; Celia Engelson, NP; Vijaya Soma, MD; Abdulwahab Abdulai; Ashanay Allen; Natella Aronova, NP; Philip Aziz, PharmD; Emily Beato; Samuel Bliss, PharmD; Jacqueline Callahan, RN; Ellie Carmody, MD; Amanda Dontino, BS; Aimee Edwin, RN; Shelby Goins; Sarah Haiken; Ramin Herati, MD; Abdonnie Holder; Janice Hong; Trishala Karmacharya; Manpreet Kaur, PharmD; Hye-Youn Kim; Alexander McMeeking, MD; Mark Mulligan, MD; Wai Ng; Edward Nirenberg; Irma Noriega, NP; Samuel Nweke; Lalitha Parameswaran, MD; Levonne Phillip, MPH; Stephanie Rettig, MPH; Marie Samanovic-Golden, PhD; Madalyn Saporito; Pamela Suman; Meron Tasissa; Michael Tuen; Julia Wagner, MPH; James Wilson; Doris Wong, PharmD; Grace Yip, BS; Samantha Yip, RN; Heekoung Youn, RN; Lisa Zhao

**University of Rochester VTEU, Rochester, NY**

Angela R. Branche, MD; Ann R. Falsey, MD; Edward E. Walsh, MD; Patrick Kingsley, BS; Arthur Zemanek, BSN, MS; Katherine Elena, BSN; Spencer Obrecht, BSN; Ian Shannon, BSN; Amy Kaychalo, BS, MS; Erin Nowicki; Sharon Moorehead; Kari Steinmetz, BA; Doreen Francis, RN; Tanya Smith, BS; William Hamilton, BS; Jeanne Holden-Wiltse, MPH, MBA; Christopher Lane, MS; Michael Peasley, BS; Samuel Diehl, BS; Kyle Richards, PharmD; Stephen Bean, PharmD; Nicole Dornbush, PharmD; Carol Cole, PharmD

**Saint Louis University, St. Louis, MO**

Sharon E. Frey, MD; Getahun Abate, MD, PhD; Zacharoula Oikonomopoulou, MD; Daniel F. Hoft, PhD, MD; Irene Graham, MD; Azra Blazevic, DVM, MPH; Tamara Blevins, MS; Kathleen Chirco, BSN; Sabrina M. DiPiazza, BSN, MA; Stanley Doublin; Heather Hoertel Douds, MSNS, BSN; Carol G. Duane, PhD, RN; Eric Eggemeyer, BA; Linda M. Eggemeyer-Sharpe, BSN; Lauren Nicole Foreman, BSN; Sarah Louise George, MD; Geoffrey J. Gorse, MD; Michelle Harris, PharmD; Helay Hassas, PharmD; Rong Hou, MD; Ryan Clark Kerr, BSN; Kate Elizabeth Liefer, BSN; Melissa J. Loyet, RN; Lainey Mejia-Jauregui, BS; Keith Meyer, BS; Tracy Renee Montauk, BSN; Karla J. Mosby, RN; Amanda Nethington, BS; Huan Ning, MD; Nicole Purcell; Joan M. Siegner, BSN, MA; Janice M. Tennant, BSN, MPH; Mei Xia, PhD; Kiana Wilder, BA; Yinyi Yu, BS; Cassandra Nicole Zehenny, BSN

**University of Texas Medical Branch, League City, TX**

Richard Rupp, MD; Laura Porterfield, MD; Amber Stanford, PA-C; Robert Cox, RN; Kristin Pollock, RN; Diane Barrett, MS; Gerrianne Casey, RN; Amy McMahan, LVN; Cori Burkett, PA-C; Essie Cox

**NYU VTEU Long Island Research Clinic at NYU Long Island School of Medicine, Mineola, NY**

Martín Bäcker, MD; Kimberly Byrnes, RN; Asif Noor, MD; Andrew B. Fleming, MD; Sigridh A. Muñoz-Gómez, MD; Diana Badillo, MD; Steven E. Carsons, MD; Sajumon K. Joseph, FNP; Sarah J. Pastolero, RN; Sophie Danziger; Monica Benitez; Maung Aung; Louis Ragolia, PhD; Alicia Vasile, RPh; April Correll, RPh; Christopher Hall; Thomas Palaia; Miloni H Thakker, MD; Lavern Harvey; Lisa Zhao

**University of Illinois at Chicago-Project WISH, Chicago, IL**

Richard M. Novak, MD; Benjamin G. Ladner, MD; Andrea Wendrow, RPh; Jesica Herrick, MD; Alfredo J. Mena Lora, MD; Scott A. Borgetti, MD; Diana L Bahena, APRN; Regina Harden, BA; Renyce Powell; David C. M. Chan, PharmD; Rebeca F. Gasari, PharmD; Michael Pacini, PharmD; Margarita M. Villarreal, CPhT; Rodrigo Reyes, ADN; Samuel M. Rene, MPH; Shannon M Whitted, BSN; Habiba Sultana, MBBS; Nanu Kunwar, BS; Tasmin Sultana, MBBS; Md R. Amin, PhD; Mahmood Ghassemi, PhD, Liam Morrissy, BS; Nia O’Neal, BS; Chasity Serrano, BS; Charlie Peterson, BA

**Duke Human Vaccine Institute, Duke University School of Medicine, Durham, NC**

Emmanuel B. Walter MD, MPH; Michael J. Smith MD, MSCE; M. Anthony Moody, MD; Kenneth E. Schmader, MD; Susan Doyle; Lynn S Harrington BSN; Lori Hendrickson BSN; Amy O’Berry MSN; Sherry Huber BSN; Janet Wootton RN, RSCN; Kelly Clark BA; Lani Banez; Stephanie Smith BA; Byron Hauser BS; Ally Odom BA; Emily Randolph BA; Krystina Yoder BA; Kathlene Chmielewski; Luis Ballon BA; Aubree Latorre; Breana Montgomery; Antony Tritz MS; Thad Gurley, MS; Margaret Pendzich

**Kaiser Permanente Washington Health Research Institute, Seattle, WA**

Lisa A. Jackson, MD, MPH; Maya Dunstan, MS, RN; Rebecca Lau, PharmD; Barbara Carste, MPH; Wesley A. Andersen, RPh, MHA, MA; Lee Barr, RN; Cassandra Bryant, BS; Joe Choe, BS; Lynn Gross, PA-C; Erika Kiniry, MPH; Bonnie Y Lam, PharmD; De Vona Lang; Stella Lee, BA; Paula J Lins, PA-C, MPH; Amy Mohelnitzky, PA-C; Marilyn Nguyen, BS; Matthew Nguyen, MPH; Melissa Resendiz Rivas, BA; Melissa Boothe Scheer, PA-C; Janice Suyehira, MD; Stacie Wellwood, LPN; Maryann K Woodford, PA-C

Susan J. Little, MD; Thomas C.S. Martin, MD; Nicole Carter, MPH; Steven Hendrickx, RN; Ajay Bharti, MD; Alyssa Phillips; Aurora Verduzco Gonzalez, NP; Cheryl Dullano; Chris Houston; Dawn Rosenblum, RN; DeeDee Pacheco; DeLys Brooks; Fang Wan; Helene Le, CPhiT; JC Alcantar; Jill Blumenthal, MD; Joseph Lencioni, MABMH; Kory Hess; Letty Muttera, PharmD; Marlene Arredondo; Megan Smyth; Megan Taylor; Melinda Stafford, PharmD; Michelle Orsburn, MD; Michelle Truong; Niamh Higgins, PharmD, MSc, AAHIVP; Nimish Patel, PharmD, PhD, AAHIVP; Rebecca Gonzalez; Vivian Maldonado

**Morehouse School of Medicine, Atlanta, GA**

Lilly C. Immergluck, MD, MS; Erica Johnson, PhD; Austin Chan, MD; Fatima Ali, MPH; Sonja Jackson; Noor Mohamed, PharmD; LaKesha Tables, MD, MPH; Norberto Fas, MD; Kay Woodson, PharmD; Saadia Khizer, MD; Jacquelyn Ali, MSA; Abdullah Warsama; Eric Gaines; Sierra Jordan Thompson; Cristina Wilson; Trisha Parker, MPH; Xiting Lin; LaTeshia Thomas Seaton, APRN; Derrick Wilson

**Howard University College of Medicine, Howard University Hospital, Washington D.C**

Siham M. Mahgoub, MD; Celia Maxwell, MD; Sarah Shami, PharmD; Edward Bauer, BS; Yuanxiu Chen, MD, PhD; Megan Ware-Pressley, MHA; Debra Ordor, RN; Linda Fletcher, RN; Emmanuel Baidoo, BS; David Jaspan, RPh, MBA; Adetokunbo Adedokun, PharmD, MPH, BCPS; Michelle Strobeck, BS; Michael A. Riga; Ashley Karen Bautista, BS

Jennifer A. Whitaker, MD; Hana M. El Sahly, MD; Wendy A. Keitel, MD; C. Mary Healy, MD; Robert L. Atmar, MD; Pedro A. Piedra, MD; Jesus Banay; Kathy Bosworth; Janet Brown, RPh; Kayla Burrell; Jeremy Castro; Tykel Eddy; Marcena Eubanks; Cathy Faw, RPh; Rachel Froebe; Alix Halter, RN; Janey John, MSN, APRN, FNP-C; Chanei Henry, AAS; Vanessa Martinez; Carol Mundell, RN; Brandie Phillips, RN; Alicia Prevost-Barthe, RN; Connie Rangel, RN; Yolanda Rayford, MS; Yvette Rugeley; Maria Shlyapobersky; Tina Sierra; Elizabeth Silguero; Lisreina Toro; Dawn Turner, RN; Chianti Wade-Bowers, RN; Jessica Woods, RN

Tara M. Babu, MD, MSCI; Anna Wald, MD, MPH; Taylor Krause, BA; Kirsten Hauge, MPH; Jina Taub, ARNP; Dana Varon, ARNP; Britt Murphy, ARNP; Morissa Pertik, PA-C; T. Nui Pholsena, ARNP; Alyssa Braun, BS; Mark Drummond, BS; Jessica Heimonen, MPH; Amy Link, BS; Lindsey McClellan, BS; Jessica Moreno, BS; Chloe Wilkens, BS; Matt Seymour, MPH; Lawrence Hemingway, BS; Jean Mernaugh, BS; Chris McClurkan, BS; Kerry Laing, PhD; Meredith Potochnic, PharmD; Joong Kim, PharmD; Bao-Chao Vo, PhT

**University of Alabama at Birmingham, Birmingham, AL**

Paul A. Goepfert, MD; Jenna Weber, RN; Savannah Spaulding, RN; Heather Logan, CRNP; Faye Heard; Foreamben Patel; Michelle Chambers

**Tulane University School of Medicine, New Orleans, LA**

Dahlene N. Fusco, MD; Arnaud C. Drouin, MD; Florice K. Numbi, MD; Hamada F. Rady, PhD; Crystal A. Ward, MSN; Quinn M. Powers, MS; William E. Casey, BS; Brian P. Logarbo, MD; Shae P. Williams, BS; Emily Callegari, MSN

**IDCRC Principal Investigators**

David S. Stephens, MD; Kathleen M. Neuzil, MD

**IDCRC Leadership Operations Center**

Monica M. Farley, MD; Jeanne Marrazzo, MD; Sidnee Paschal Young

**IDCRC Clinical Operations Unit**

Jeffery Lennox, MD; Robert L. Atmar, MD; Linda McNeil FHI360

**IDCRC Laboratory Operations Unit – Fred Hutchinson Cancer Center and University of Washington, Seattle, WA**

Christine M. Posavad, PhD; Megan A. Meagher, BS; Michael Stirewalt, MBA; John Hural, PhD; Weston Lawler, BA; Lexi Tanser, MA; Julie McElrath, MD, PhD; Mike Gale, PhD

**IDCRC Statistical and Data Science Unit**

Elizabeth Brown, PhD

**University of Maryland**

Kirsten E. Lyke, MD

**FHI360, Durham, NC**

Kuleni Abebe, MSc

**The Emmes Company, LLC, Rockville, MD**

Teri C. Lewis, BS; Lisa A. Giebeig, MS; Theresa M. Engel, MFS.; Caleb J. Griffith, MPH; Wendi L. McDonald, BSN; Alissa E. Burkey, MS; Lisa B. Hoopengardner, MS; Jessica E. Linton, MS; Nikki L. Gettinger, MPH; Aroussiak Bowen; Beth R. Baseler, MS; Vanessa S. Eccard-Koons, MS; Charles W. R. Hofsommer, JD; Thomas C. Sova, JD; Gary A. Krauss

David C Montefiori, PhD; Amanda Eaton, MBA; Francesca Suman, MS.

**Smith’s Laboratory, Cambridge, UK**

Derek J Smith, PhD; Antonia Netzl; Samuel H Wilks, PhD; Sina Türeli, PhD; Ana Mosterín Höpping, PhD; Samuel Turner; Sarah James, MD; Poppy Roth

**Division of Microbiology and Infectious Diseases, National Institute of Allergy and Infectious Diseases, National Institutes of Health, Bethesda, MD.**

Marina Lee, PhD; Mamodikoe Makhene, MD; Mohamed Elsafy, MD; Rhonda Pikaart-Tautges, BS; Janice Arega, MS: Binh Hoang, RPh; Dan Curtin; Hyung Koo, BSN; Elisa Sindall, BSN; Aya Nakamura, RN, MS; Audria Crowder, BS; Guinevere Chun, RN, BSN, MSHS; Frank Kenny, PhD MPH; Seemi Patel, RHP, PharmD; Sonia Gales, MS; Ahsen Khan, JD; Walla Dempsey, PhD; Robert Jurao-RN, BSN; Sonja Crandon, BSN; Seema U. Nayak, MD; Marciela M DeGrace, PhD; Diane J Post, PhD; Paul C Roberts, PhD; John H Beigel, MD; SAVE Program

---

**TABLE S1.**
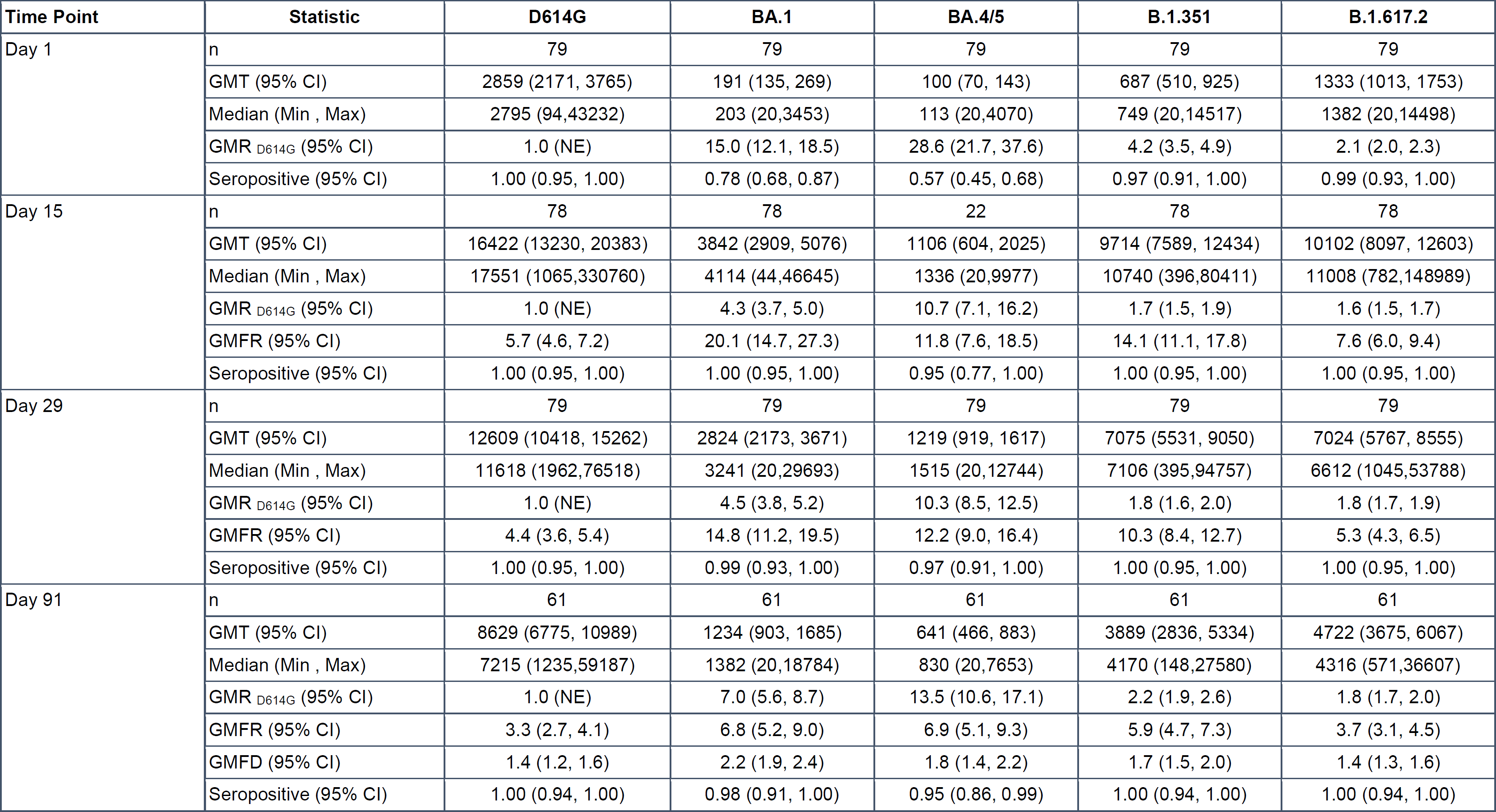
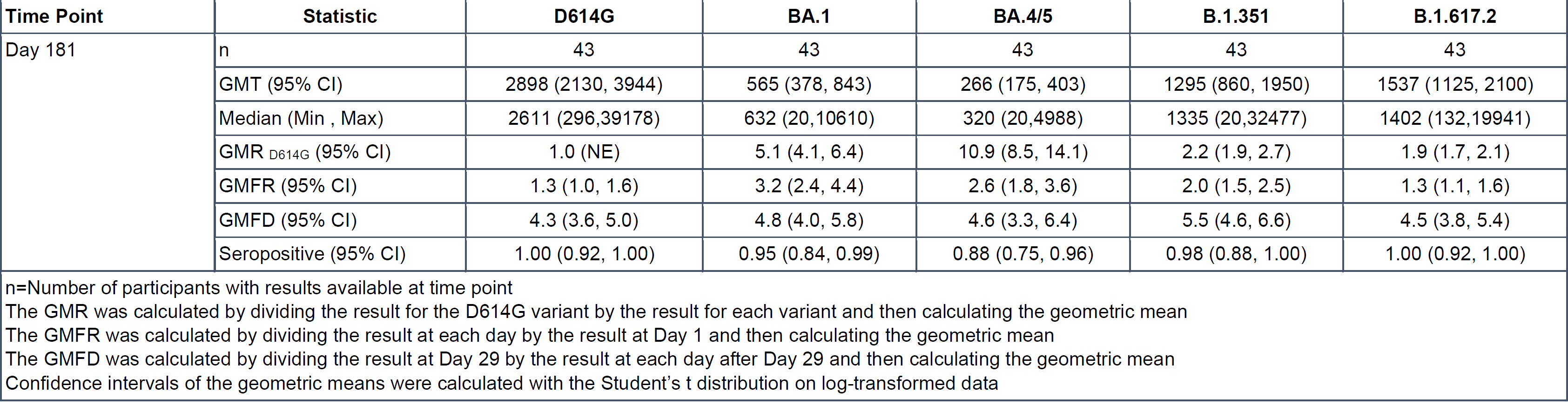
Pseudovirus Neutralization Assay Summary Results after vaccination with 1 dose of Moderna mRNA-1273 Beta + Omicron BA.1, for participants without a history of prior infection, against variants of concern at baseline (single dose) and Days 15, 29, 91, 181.

**TABLE S2.**
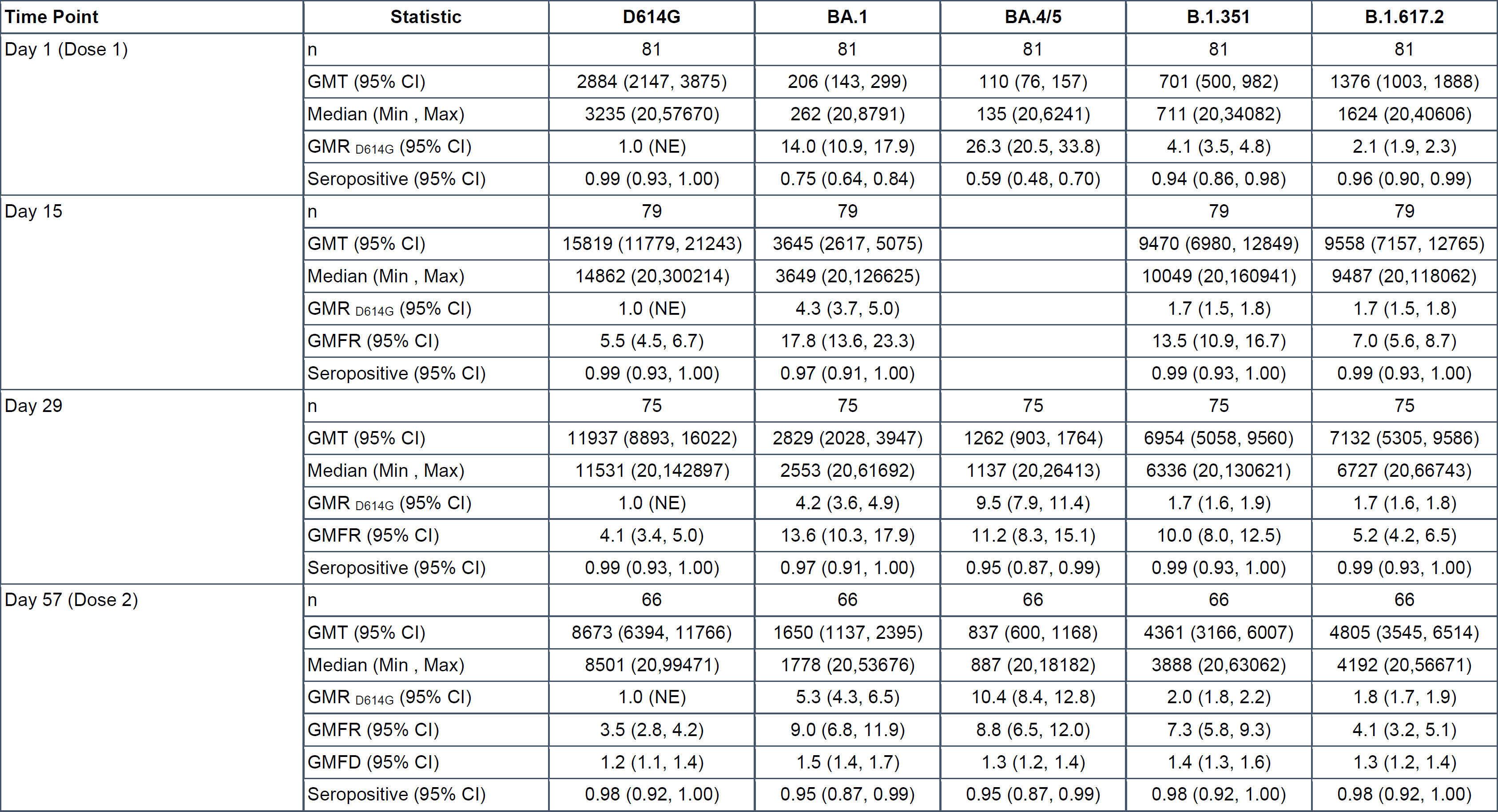
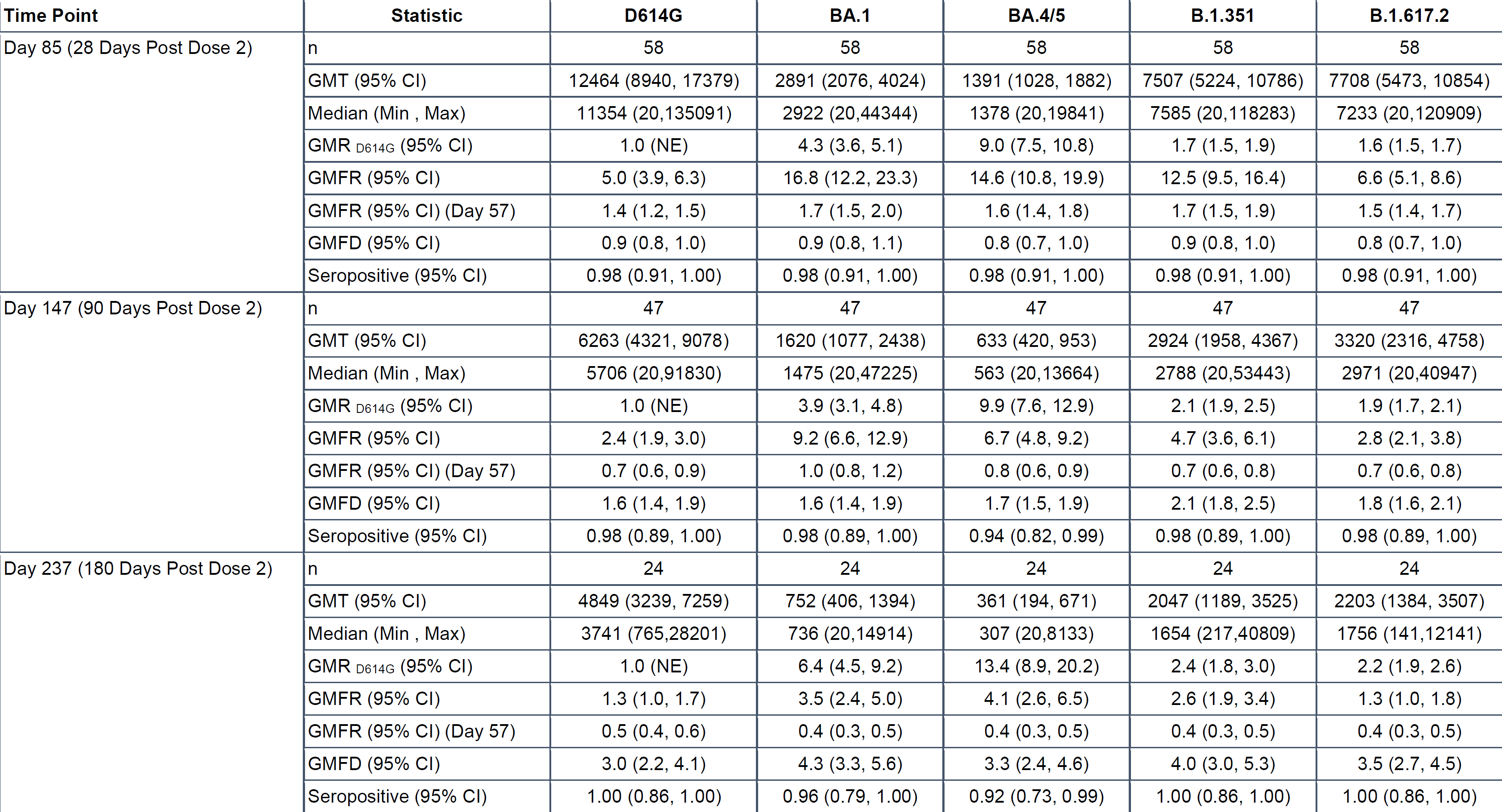
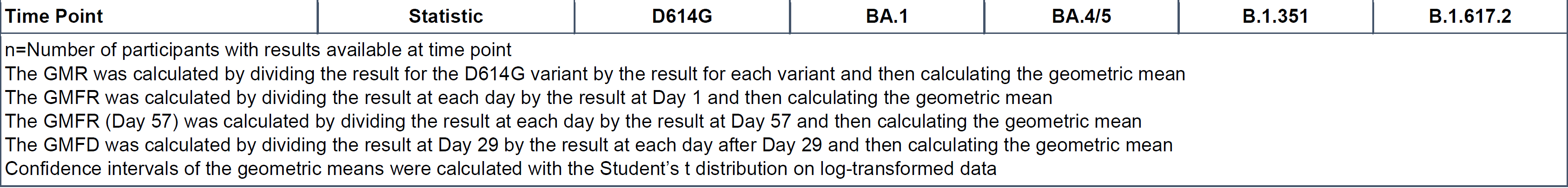
Pseudovirus Neutralization Assay Summary Results after vaccination with 2 doses of Moderna mRNA-1273 Beta + Omicron BA.1, for participants without a history of prior infection at baseline, against variants of concern at baseline (first dose) and Days 15, 29, 57 (second dose), 85, 147 and 237.

**TABLE S3.**
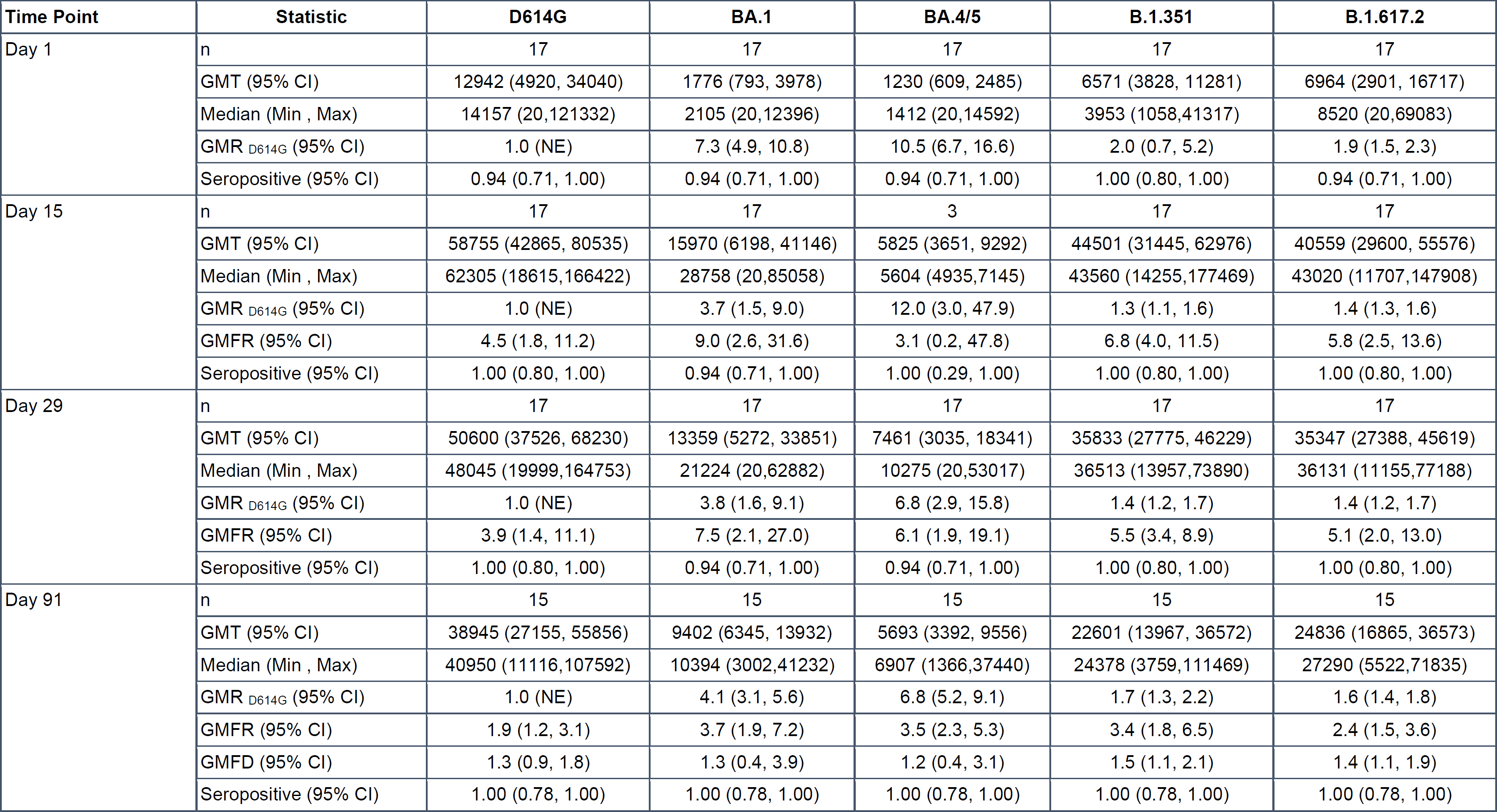
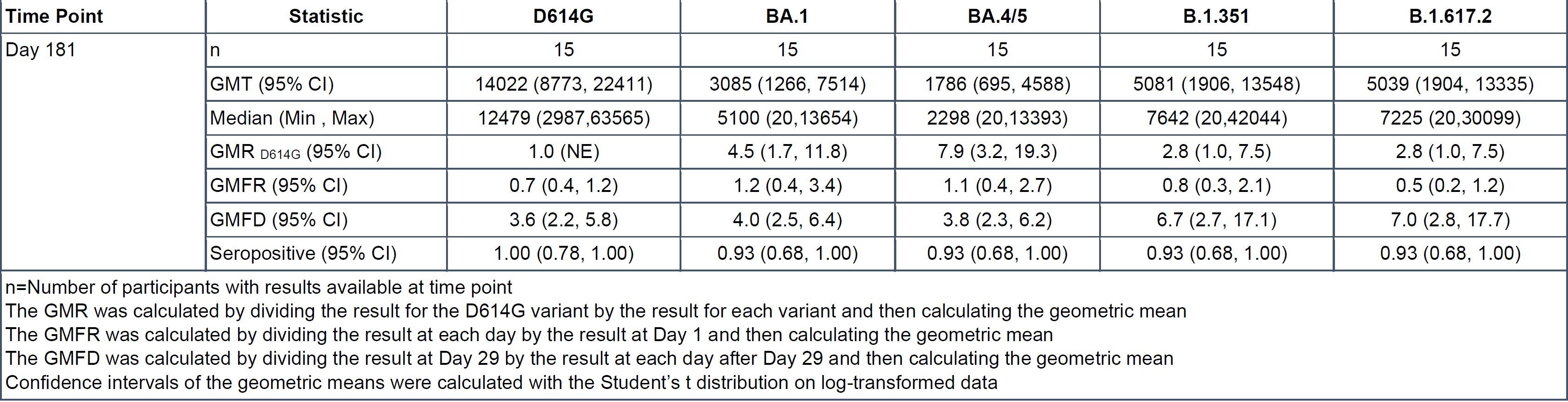
Pseudovirus Neutralization Assay Summary Results after vaccination with 1 dose of Moderna mRNA-1273 Beta + Omicron BA.1, for participants with a history of prior infection, against variants of concern at baseline (single dose) and Days 15, 29, 91, 181.

**TABLE S4.** Pseudovirus Neutralization Assay Summary Results after vaccination with 2 doses of Moderna mRNA-1273 Beta + Omicron BA.1, for participants with a history of prior infection, against variants of concern at baseline (first dose) and Days 15, 29, 57 (second dose), 85, 147 and 237.

| Time Point | Statistic | D614G | BA.1 | BA.4/5 | B.1.351 | B.1.617.2 |
| --- | --- | --- | --- | --- | --- | --- |
| Day 1 (Dose 1) | n | 21 | 21 | 21 | 21 | 21 |
|  | GMT (95% CI) | 9522 (5381, 16851) | 1079 (404, 2878) | 553 (228, 1344) | 3085 (1204, 7905) | 3576 (1395, 9171) |
|  | Median (Min , Max) | 13237 (381,76328) | 2362 (20,15942) | 838 (20,7823) | 3962 (20,34071) | 6758 (20,43817) |
|  | GMR<sub>D614G</sub> (95% CI) | 1.0 (NE) | 8.8 (4.7, 16.6) | 17.2 (9.2, 32.1) | 3.1 (1.6, 5.9) | 2.7 (1.4, 5.1) |
|  | Seropositive (95% CI) | 1.00 (0.84, 1.00) | 0.86 (0.64, 0.97) | 0.81 (0.58, 0.95) | 0.90 (0.70, 0.99) | 0.90 (0.70, 0.99) |
| Day 15 | n | 21 | 21 |  | 21 | 21 |
|  | GMT (95% CI) | 37934 (22711, 63361) | 11876 (4820, 29257) |  | 30175 (17248, 52791) | 17948 (7538, 42733) |
|  | Median (Min , Max) | 47039 (1800,208690) | 18447 (20,94116) |  | 44277 (789,131933) | 32463 (20,148746) |
|  | GMR<sub>D614G</sub> (95% CI) | 1.0 (NE) | 3.2 (1.6, 6.5) |  | 1.3 (1.1, 1.5) | 2.1 (1.0, 4.3) |
|  | GMFR (95% CI) | 4.0 (2.9, 5.5) | 11.0 (6.8, 17.7) |  | 9.8 (4.8, 20.0) | 5.0 (3.1, 8.1) |
|  | Seropositive (95% CI) | 1.00 (0.84, 1.00) | 0.95 (0.76, 1.00) |  | 1.00 (0.84, 1.00) | 0.95 (0.76, 1.00) |
| Day 29 | n | 20 | 20 | 20 | 20 | 20 |
|  | GMT (95% CI) | 21358 (8873, 51410) | 7898 (3295, 18931) | 3656 (1628, 8209) | 15433 (6486, 36725) | 12091 (5246, 27868) |
|  | Median (Min , Max) | 28826 (20,143634) | 11742 (20,72318) | 4723 (20,32606) | 21894 (20,136949) | 16764 (20,134673) |
|  | GMR<sub>D614G</sub> (95% CI) | 1.0 (NE) | 2.7 (2.0, 3.7) | 5.8 (4.0, 8.6) | 1.4 (1.2, 1.7) | 1.8 (1.5, 2.1) |
|  | GMFR (95% CI) | 1.9 (0.8, 4.5) | 6.0 (3.8, 9.5) | 5.6 (3.5, 8.9) | 3.9 (2.6, 5.9) | 2.6 (1.8, 3.8) |
|  | Seropositive (95% CI) | 0.95 (0.75, 1.00) | 0.95 (0.75, 1.00) | 0.95 (0.75, 1.00) | 0.95 (0.75, 1.00) | 0.95 (0.75, 1.00) |
| Day 57 (Dose 2) | n | 15 | 15 | 15 | 15 | 15 |
|  | GMT (95% CI) | 23074 (12633, 42145) | 7412 (3093, 17758) | 3002 (1342, 6712) | 17783 (9291, 34035) | 14015 (7690, 25542) |
|  | Median (Min , Max) | 26445 (1929,119671) | 7660 (152,72907) | 3137 (113,33825) | 24110 (1678,128140) | 11773 (1301,100276) |
|  | GMR<sub>D614G</sub> (95% CI) | 1.0 (NE) | 3.1 (1.9, 5.0) | 7.7 (4.7, 12.6) | 1.3 (1.0, 1.6) | 1.6 (1.4, 1.9) |
|  | GMFR (95% CI) | 2.1 (1.5, 3.1) | 7.1 (3.3, 15.4) | 5.5 (2.8, 10.8) | 5.2 (2.4, 11.4) | 3.6 (1.6, 8.0) |
|  | GMFD (95% CI) | 0.9 (0.3, 2.3) | 0.9 (0.4, 2.1) | 1.0 (0.5, 2.2) | 0.8 (0.3, 2.1) | 0.8 (0.3, 1.9) |
|  | Seropositive (95% CI) | 1.00 (0.78, 1.00) | 1.00 (0.78, 1.00) | 1.00 (0.78, 1.00) | 1.00 (0.78, 1.00) | 1.00 (0.78, 1.00) |
| Day 85 (28 Days Post Dose 2) | n | 15 | 15 | 15 | 15 | 15 |
|  | GMT (95% CI) | 29718 (17259, 51171) | 11089 (5239, 23474) | 4574 (2236, 9357) | 20792 (11617, 37212) | 17052 (9726, 29895) |
|  | Median (Min , Max) | 27914 (3093,151167) | 12752 (546,68721) | 4445 (205,30021) | 20585 (2676,139245) | 16780 (1675,76860) |
|  | GMR <sub>D614G</sub> (95% CI) | 1.0 (NE) | 2.7 (1.9, 3.8) | 6.5 (4.4, 9.5) | 1.4 (1.1, 1.8) | 1.7 (1.5, 2.0) |
|  | GMFR (95% CI) | 2.7 (1.7, 4.3) | 10.6 (4.4, 25.5) | 8.4 (3.8, 18.5) | 6.1 (2.6, 14.3) | 4.3 (1.8, 10.7) |
|  | GMFR (95% CI) (Day 57) | 1.3 (1.0, 1.6) | 1.5 (1.1, 2.1) | 1.5 (1.2, 2.0) | 1.2 (1.0, 1.4) | 1.2 (1.0, 1.5) |
|  | GMFD (95% CI) | 0.7 (0.2, 1.9) | 0.6 (0.2, 1.7) | 0.7 (0.3, 1.6) | 0.7 (0.3, 1.9) | 0.7 (0.2, 1.7) |
|  | Seropositive (95% CI) | 1.00 (0.78, 1.00) | 1.00 (0.78, 1.00) | 1.00 (0.78, 1.00) | 1.00 (0.78, 1.00) | 1.00 (0.78, 1.00) |
| Day 147 (90 Days Post Dose 2) | n | 15 | 15 | 15 | 15 | 15 |
|  | GMT (95% CI) | 13141 (7990, 21613) | 3612 (1105, 11803) | 1262 (405, 3932) | 9284 (5088, 16940) | 4805 (1751, 13187) |
|  | Median (Min , Max) | 13750 (1732,59538) | 4552 (20,62571) | 2288 (20,20727) | 10118 (988,49230) | 7201 (20,39264) |
|  | GMR <sub>D614G</sub> (95% CI) | 1.0 (NE) | 3.6 (1.4, 9.3) | 10.4 (4.4, 24.9) | 1.4 (1.1, 1.8) | 2.7 (1.1, 6.6) |
|  | GMFR (95% CI) | 1.2 (0.8, 1.8) | 3.5 (2.0, 5.9) | 2.3 (1.3, 4.1) | 2.7 (1.0, 7.4) | 1.2 (0.8, 1.9) |
|  | GMFR (95% CI) (Day 57) | 0.6 (0.4, 0.7) | 0.5 (0.2, 1.0) | 0.4 (0.2, 0.8) | 0.5 (0.4, 0.7) | 0.3 (0.2, 0.7) |
|  | GMFD (95% CI) | 1.5 (0.5, 4.5) | 1.9 (1.4, 2.6) | 2.4 (1.6, 3.7) | 1.6 (0.5, 4.9) | 2.3 (1.8, 2.9) |
|  | Seropositive (95% CI) | 1.00 (0.78, 1.00) | 0.93 (0.68, 1.00) | 0.87 (0.60, 0.98) | 1.00 (0.78, 1.00) | 0.93 (0.68, 1.00) |
| Day 237 (180 Days Post Dose 2) | n | 10 | 10 | 10 | 10 | 10 |
|  | GMT (95% CI) | 11790 (4965, 27998) | 1240 (240, 6390) | 730 (182, 2927) | 3399 (615, 18777) | 2663 (579, 12246) |
|  | Median (Min , Max) | 12764 (1208,94288) | 2534 (20,32573) | 1601 (20,8973) | 6727 (20,89710) | 3984 (20,30180) |
|  | GMR <sub>D614G</sub> (95% CI) | 1.0 (NE) | 9.5 (2.7, 32.9) | 16.1 (5.5, 47.8) | 3.5 (0.8, 14.3) | 4.4 (1.2, 16.4) |
|  | GMFR (95% CI) | 1.0 (0.6, 1.6) | 1.4 (0.7, 2.8) | 1.6 (0.7, 3.5) | 1.3 (0.7, 2.4) | 0.8 (0.5, 1.3) |
|  | GMFR (95% CI) (Day 57) | 0.5 (0.4, 0.8) | 0.2 (0.1, 0.5) | 0.3 (0.1, 0.7) | 0.2 (0.1, 0.8) | 0.2 (0.1, 0.6) |
|  | GMFD (95% CI) | 1.2 (0.2, 6.8) | 4.0 (2.4, 6.7) | 3.2 (1.9, 5.4) | 3.0 (1.9, 4.6) | 3.2 (2.1, 4.6) |
|  | Seropositive (95% CI) | 1.00 (0.69, 1.00) | 0.90 (0.55, 1.00) | 0.90 (0.55, 1.00) | 0.90 (0.55, 1.00) | 0.90 (0.55, 1.00) |
| n=Number of participants with results available at time point<br>The GMR was calculated by dividing the result for the D614G variant by the result for each variant and then calculating the geometric mean<br>The GMFR was calculated by dividing the result at each day by the result at Day 1 and then calculating the geometric mean<br>The GMFR (Day 57) was calculated by dividing the result at each day by the result at Day 57 and then calculating the geometric mean<br>The GMFD was calculated by dividing the result at Day 29 by the result at each day after Day 29 and then calculating the geometric mean<br>Confidence intervals of the geometric means were calculated with the Student's t distribution on log-transformed data |  |  |  |  |  |  |

**TABLE S5.** List of COVAIL US Sites **Site name** George Washington University University of Rochester Medical Center Hope Clinic of the Emory Vaccine Center Brigham and Women’s Hospital, Harvard Medical School Saint Louis University Baylor College of Medicine University of California San Diego Emory Center for Childhood Infections and Vaccines of Children’s Healthcare of Atlanta Duke Human Vaccine Institute, Duke University School of Medicine University of Illinois at Chicago, Project WISH University of Texas Medical Branch Kaiser Permanente Washington Health Research Institute University of Washington New York University Manhattan Research Clinic, NYU Grossman School of Medicine Zuckerberg San Francisco General Hospital, University of California at San Francisco Morehouse School of Medicine Washington University School of Medicine NYU Long Island Research Clinic, NYU Long Island School of Medicine University of Iowa College of Medicine Howard University Hospital, Howard University Hospital University of Alabama at Birmingham

**TABLE S6.** Eligible Regimens for Primary Vaccination Series and Boost

| **Primary series**<br>**vaccine**<br>**manufacturer** | **Number of**<br>**doses in**<br>**primary**<br>**series** | **Primary**<br>**series**<br>**dose** | **Interval**<br>**between 1st**<br>**and 2nd**<br>**dose** | **Number of**<br>**booster**<br>**doses** | **Booster**<br>**dose** | **Allowed interval**<br>**between primary**<br>**series and booster**<br>**dose\*** |
| --- | --- | --- | --- | --- | --- | --- |
| Pfizer BioNTech | 2 | 30 µg | At least 17<br>days | 1 | 30 mcg | ≥ 5 months |
| Moderna | 2 | 100 µg | At least 24<br>days | 1 | 50 mcg | ≥ 5 months |
| Janssen | 1 | 5×10<sup>10</sup> viral<br>particles | Not applicable | 1 | 5×10<sup>10</sup> viral<br>particles | ≥ 2 months |

\*homologous or heterologous boosters were acceptable

**TABLE S7:**
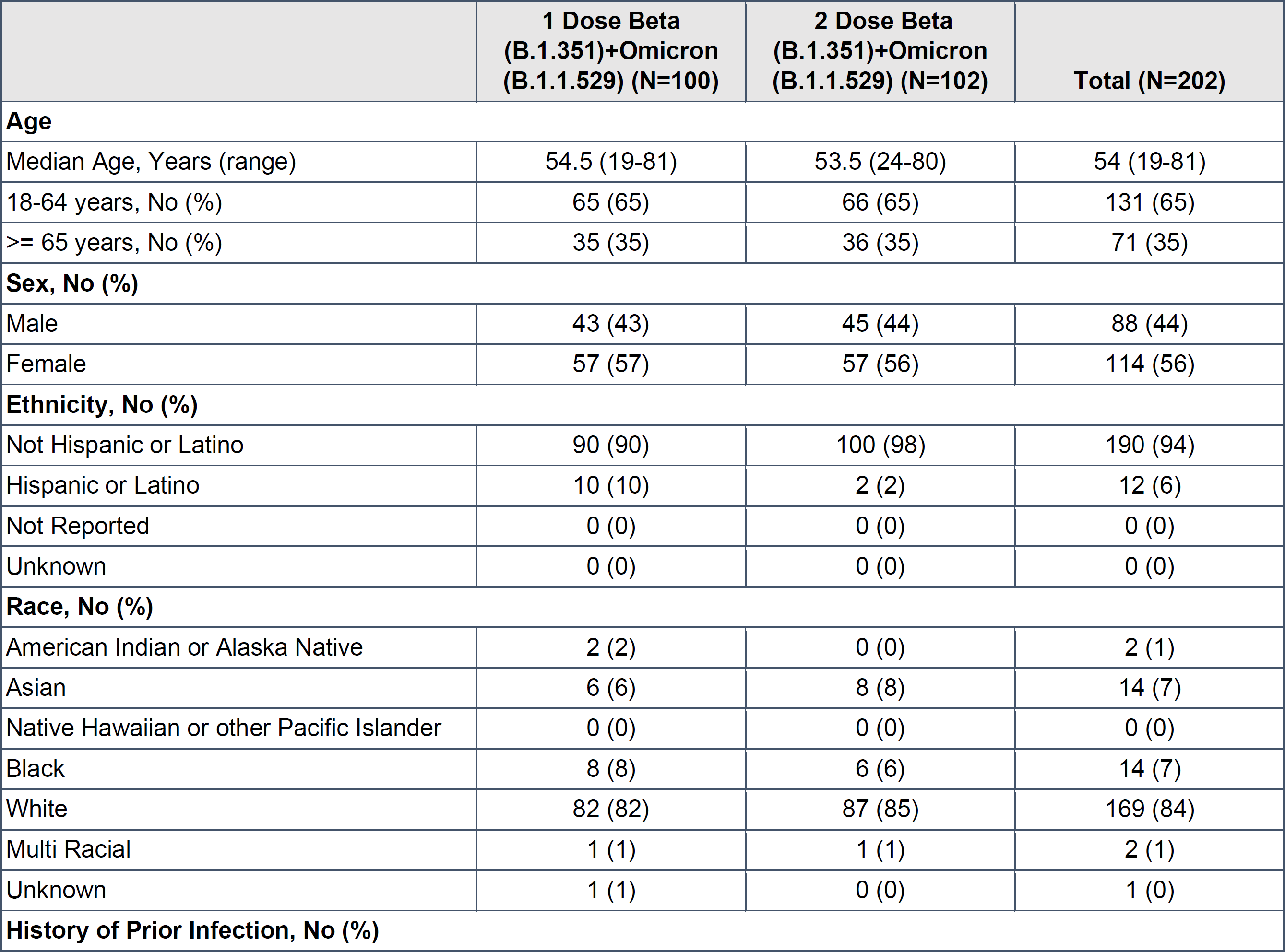
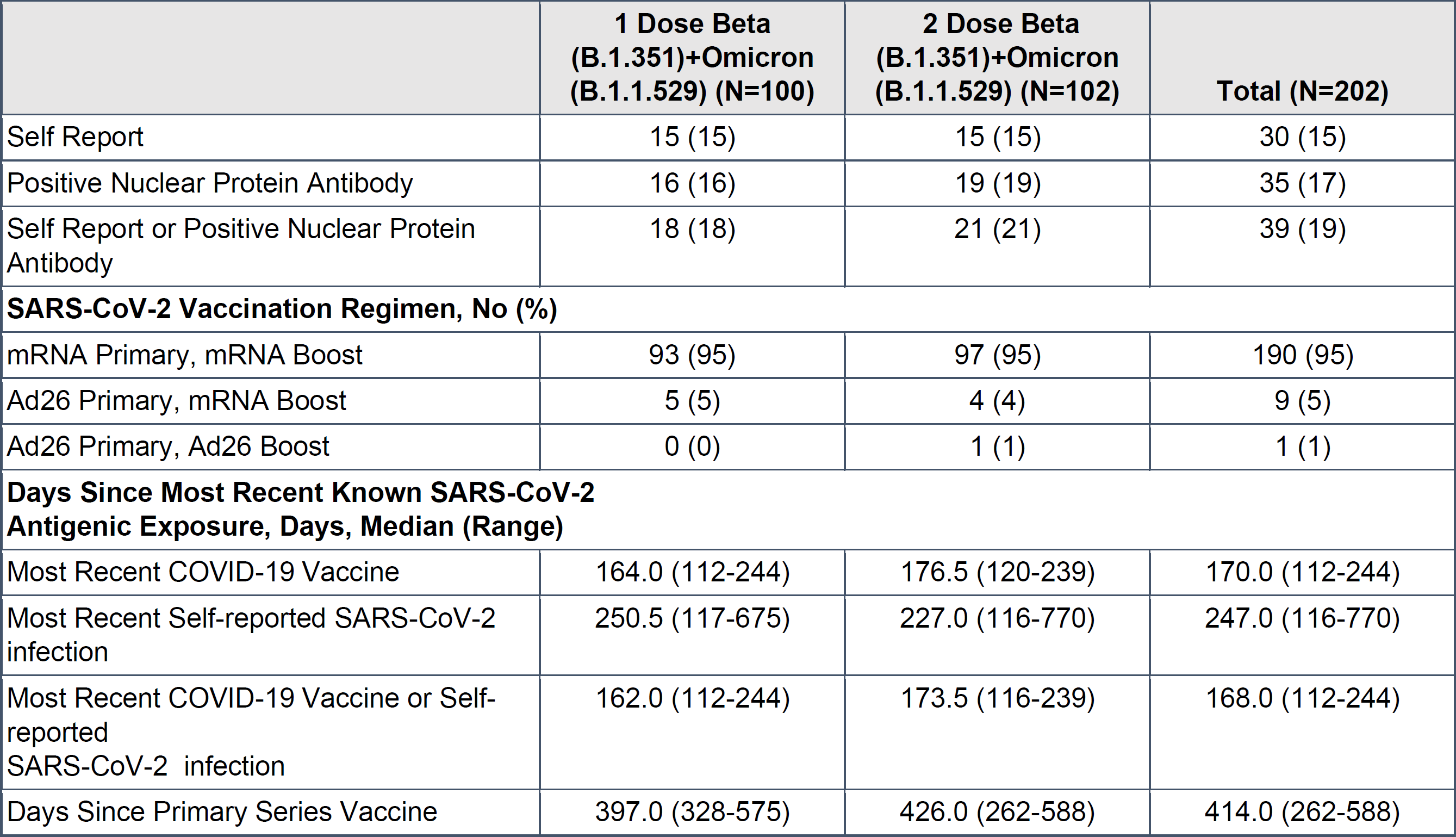
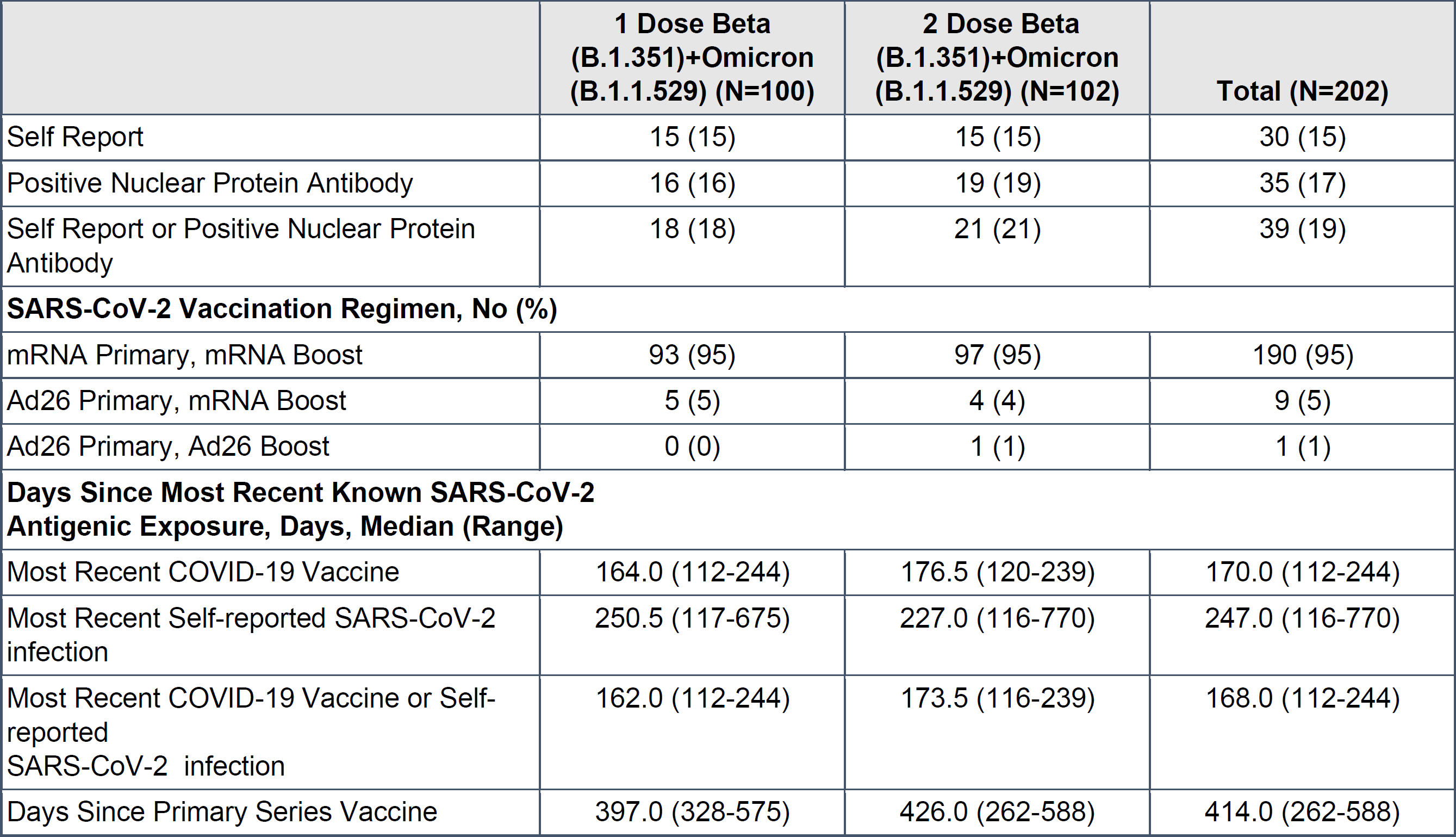
Summary of Demographic and Baseline Characteristics by Vaccination Group

|  | **1 Dose Beta**<br>**(B.1.351)+Omicron**<br>**(B.1.1.529) (N=100)** | **2 Dose Beta**<br>**(B.1.351)+Omicron**<br>**(B.1.1.529) (N=102)** | **Total (N=202)** |
| --- | --- | --- | --- |
| **Age** |  |  |  |
| Median Age, Years (range) | 54.5 (19-81) | 53.5 (24-80) | 54 (19-81) |
| 18-64 years, No (%) | 65 (65) | 66 (65) | 131 (65) |
| >= 65 years, No (%) | 35 (35) | 36 (35) | 71 (35) |
| **Sex, No (%)** |  |  |  |
| Male | 43 (43) | 45 (44) | 88 (44) |
| Female | 57 (57) | 57 (56) | 114 (56) |
| **Ethnicity, No (%)** |  |  |  |
| Not Hispanic or Latino | 90 (90) | 100 (98) | 190 (94) |
| Hispanic or Latino | 10 (10) | 2 (2) | 12 (6) |
| Not Reported | 0 (0) | 0 (0) | 0 (0) |
| Unknown | 0 (0) | 0 (0) | 0 (0) |
| **Race, No (%)** |  |  |  |
| American Indian or Alaska Native | 2 (2) | 0 (0) | 2 (1) |
| Asian | 6 (6) | 8 (8) | 14 (7) |
| Native Hawaiian or other Pacific Islander | 0 (0) | 0 (0) | 0 (0) |
| Black | 8 (8) | 6 (6) | 14 (7) |
| White | 82 (82) | 87 (85) | 169 (84) |
| Multi Racial | 1 (1) | 1 (1) | 2 (1) |
| Unknown | 1 (1) | 0 (0) | 1 (0) |
| **History of Prior Infection, No (%)** |  |  |  |
| Self Report | 15 (15) | 15 (15) | 30 (15) |
| Positive Nuclear Protein Antibody | 16 (16) | 19 (19) | 35 (17) |
| Self Report or Positive Nuclear Protein Antibody | 18 (18) | 21 (21) | 39 (19) |
| **SARS-CoV-2 Vaccination Regimen, No (%)** |  |  |  |
| mRNA Primary, mRNA Boost | 93 (95) | 97 (95) | 190 (95) |
| Ad26 Primary, mRNA Boost | 5 (5) | 4 (4) | 9 (5) |
| Ad26 Primary, Ad26 Boost | 0 (0) | 1 (1) | 1 (1) |
| **Days Since Most Recent Known SARS-CoV-2 Antigenic Exposure, Days, Median (Range)** |  |  |  |
| Most Recent COVID-19 Vaccine | 164.0 (112-244) | 176.5 (120-239) | 170.0 (112-244) |
| Most Recent Self-reported SARS-CoV-2 infection | 250.5 (117-675) | 227.0 (116-770) | 247.0 (116-770) |
| Most Recent COVID-19 Vaccine or Self-reported SARS-CoV-2 infection | 162.0 (112-244) | 173.5 (116-239) | 168.0 (112-244) |
| Days Since Primary Series Vaccine | 397.0 (328-575) | 426.0 (262-588) | 414.0 (262-588) |

## References

1. Branche AR, Rouphael NG, Diemert DJ, et al. SARS-CoV-2 Variant Vaccine Boosters Trial: Preliminary Analyses. medRxiv. 2022:2022.2007.2012.22277336.

2. US Food and Drug Administration (FDA). Coronavirus (COVID-19) Update: FDA Authorizes Second Booster Dose of Two COVID-19 Vaccines for Older and Immunocompromised Individuals. 2022.

3. Huang Y, Borisov O, Kee JJ, et al. Calibration of two validated SARS-CoV-2 pseudovirus neutralization assays for COVID-19 vaccine evaluation. Scientific Reports. 2021;11(1):23921.

4. Ellebedy AH, Nachbagauer R, Jackson KJL, et al. Adjuvanted H5N1 influenza vaccine enhances both cross-reactive memory B cell and strain-specific naive B cell responses in humans. Proc Natl Acad Sci U S A. 2020;117(30):17957–17964.

## Supplemental Reference

1. Shen X, Chalkias S, Feng J, et al. Neutralization of SARS-CoV-2 Omicron BA.2.75 after mRNA-1273 Vaccination. N Engl J Med. 2022;387(13):1234-1236.

